# Early transmission of COVID-19 has an optimal temperature but late transmission decreases in warm climate

**DOI:** 10.1101/2020.05.14.20102459

**Authors:** Xinru Wan, Chaoyuan Cheng, Zhibin Zhang

**Affiliations:** State Key Laboratory of Integrated Management on Pest Insects and Rodents in Agriculture, Institute of Zoology, Chinese Academy of Sciences, Beijing 100101, China; University of Chinese Academy of Sciences, Beijing 100049, China; CAS Center for Excellence in Biotic Interactions, University of Chinese Academy of Sciences, Beijing 100049, China

**Keywords:** COVID-19, transmission ability, control efficiency, temperature, precipitation, disease prevention, control

## Abstract

The COVID-19 novel virus, as an emerging highly pathogenic agent, has caused a pandemic. Revealing the influencing factors affecting transmission of COVID-19 is essential to take effective control measures. Several previous studies suggested that the spread of COVID-19 was likely associated with temperature and/or humidity. But, a recent extensive review indicated that conclusions on associations between climate and COVID-19 were elusive with high uncertainty due to caveats in most previous studies, such as limitations in time and space, data quality and confounding factors. In this study, by using a more extensive global dataset covering 578 time series from China, USA, Europe and the rest of the world, we show that climate show distinct impacts on early and late transmission of COVID-19 in the world after excluding the confounding factors. The early transmission ability of COVID-19 peakedaround 6.3°C without or with little human intervention, but the later transmission ability was reduced in high temperature conditions under human intervention, probably driven by increased control efficiency of COVID-19. The transmission ability was positively associated with the founding population size of early reported cases and population size of a location. Our study suggested that with the coming summer seasons, the transmission risk of COVID-19 would increase in the high-latitude or high-altitude regions but decrease in low-latitude or low-altitude regions; human intervention is essential in containing the spread of COVID-19 around the world.

## INTRODUCTION

Recently, a novel coronavirus (defined as SARS-CoV-2 by the International Committee on Taxonomy of Viruses) is spreading rapidly in the world. It has caused incredible damage to public health around the world. By April 4, 2020, a total of 1,051,635 confirmed cases of COVID-19 over 200 countries or regions in the world were reported. There is an urgent need to contain the fast-expansion of COVID-19 in the world.

Revealing the influencing factors on the spread of COVID-19 is extremely important to take effective control measures. There is evidence that human movement could facilitated the spread of COVID-19 around the world (Yang et al., 2020), thus, lockdown of the epicenter, social distancing and isolation of infected patients have been widely adopted to prevent and control COVID-19 (Tian et al., 2020). However, the knowledge about the impacts of climate on the spread of COVID-19 is still limited and unclear. It is widely speculated that the COVID-19 virus could be prohibited in warmer seasons because many viral diseases show an explicit seasonality and annual cycles, such as influenza epidemics (Martinez, 2018), human coronaviruses (HCoV-229E, HCoV-HKU1, HCoV-NL63, and HCoV-OC43) (Gaunt, Hardie, Claas, Simmonds, & Templeton, 2010) and the SARS coronavirus (Chan et al., 2011; Yuan et al., 2006). Several studies suggested that the spread of COVID-19 was associated with temperature and/or humidity (Araujo & Naimi, 2020; Bannister-Tyrrell, Meyer, Faverjon, & Cameron, 2020; Chen et al., 2020; Ficetola & Rubolini, 2020; Ma et al., 2020; Shi et al., 2020; Wang, Tang, Feng, & Lv, 2020). But, a recent extensive review by the National Academy of Sciences, Engineering and Medicine of the United States of America indicated that conclusions on associations between climate and COVID-19 were elusive with high uncertainty due to significant caveats in most previous studies such as limitation in time and space, data quality and confounding factors(National Academies of Sciences & Medicine, 2020). Therefore, it is necessary to reveal the impacts of climate factors on the spread of COVID-19 by using a more extensive global dataset covering a large geographic and climatic variation, and by excluding the impacts of human factors and spatial autocorrelation.

Here, by using global data of COVID-19 cumulative cases released by WHO or national healthy committee or institutions (Fig. 1), we estimated the maximum daily increase rate (early transmission ability without human intervention), the average daily increase rate (later transmission ability with human intervention), and the regression slope of daily increase rate with cumulative cases (the control efficiency)(see Methods). We analyzed associations of these three parameters with both human factors (founding population size of the early reported patients within one week and population size of a location) and climate factors (air temperature and precipitation) in China, USA, Europe and the world. We found that both human and climate factors affected the spread of COVID-19, but temperature showed distinct impacts on the early and late spread of COVID-19.

**Figure 1.**
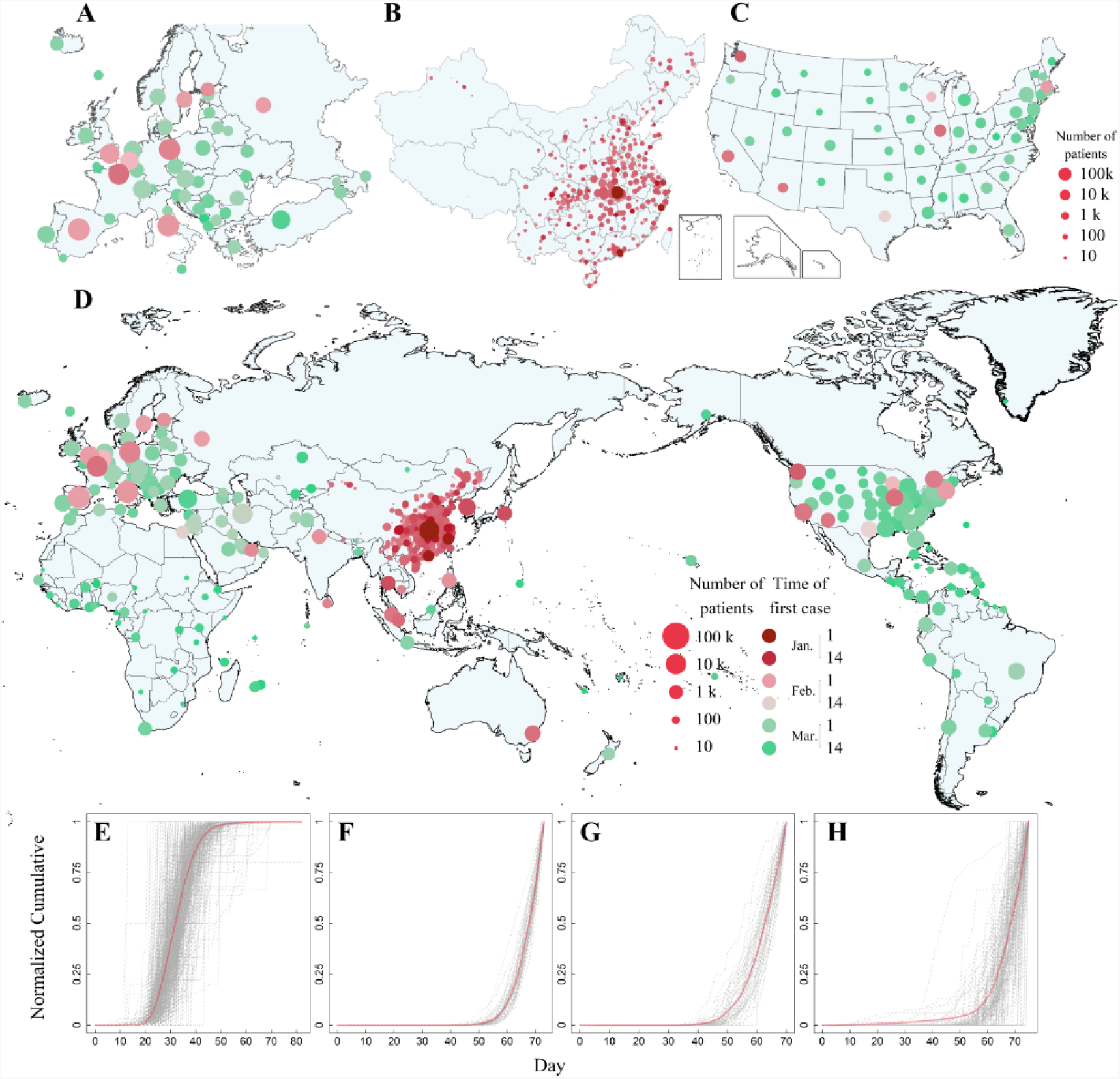
The number of cumulative cases of COVID-19 from 1 January to 4 April,2020in Europe (A), China (B), USA (C), and the world (D), and the normalized time series of cumulative cases of each location in China (E), USA (F), Europe (G), and rest of the world (H) A-D. The spread of COVID-19 around the world. The color of the circles shows the time of the first reported coronavirus case to the recipient location (number of days from 1 January; the earlier, the darker). The relative size of the circle shows the number of patients of the location represented by the capital of the country of administrative regions. E. Time series of the normalized cumulative cases of different cities or prefectures in China. F. Time series of the normalized cumulative cases of different states in USA. G. Time series of the normalized cumulative cases of various countries in Europe. H. Time of the normalized cumulative cases of other countries or regions in the world excluding China, USA and Europe. The solid red line indicates loess regression with span = 0.25. The dash grey line indicates temporal change of cumulative cases of COVID-19 for a given location. The date with first report of COVID-19 patient was set to zero.

## RESULTS AND DISCUSSION

### Impacts of climate factor

Analysis using linear generalized additive models (GAM) models indicated that air temperature showed a consistent, significant and negative association with the average daily increase rate (*r*_i_) in China, Europe and the world, the maximum daily increase rate (*a*_i_) in the world (Table 1, SI Appendix, Table S2), indicating high temperature reduced the transmission ability of COVID-19 with and without human intervention. These results are generally consistent with observation of several previous studies using different parameters of transmission severity such as incident cases (Araujo & Naimi, 2020; Bannister-Tyrrell et al., 2020; Qi et al., 2020), reproductive number (M. Wang et al., 2020) or mortality (Ma et al., 2020), except for one study (Yao et al., 2020). Using niche models, tropical climates are less vulnerable than temperate climates (Araujo & Naimi, 2020). Temperature is positively associated with COVID-19 mortality in Wuhan (Ma et al., 2020). Triplett (2020) found a downward trend of COVID-19 cases with a maximum temperature above 22.5°C (Triplett, 2020). Wang *et al*. (2020b) reported that high temperature and high humidity significantly reduced the effective reproduction number of COVID-19 in China (M. Wang et al., 2020). Qi *et al*. (2020) reported the negative association between incidence of COVID-19 and temperature or relative humidity in China (Qi et al., 2020). However, in one study, no association between the basic reproduction number of COVID-19 with temperature or UV radiation in Chinese cities was found (Yao et al., 2020). We found precipitation showed a significant and negative association with the average daily increase rate (*r*_i_) in China, suggesting a wet climate might decrease the transmission ability of COVID-19 (Table 1), which is consistent with a previous study (Ma et al., 2020).

**Table 1:**
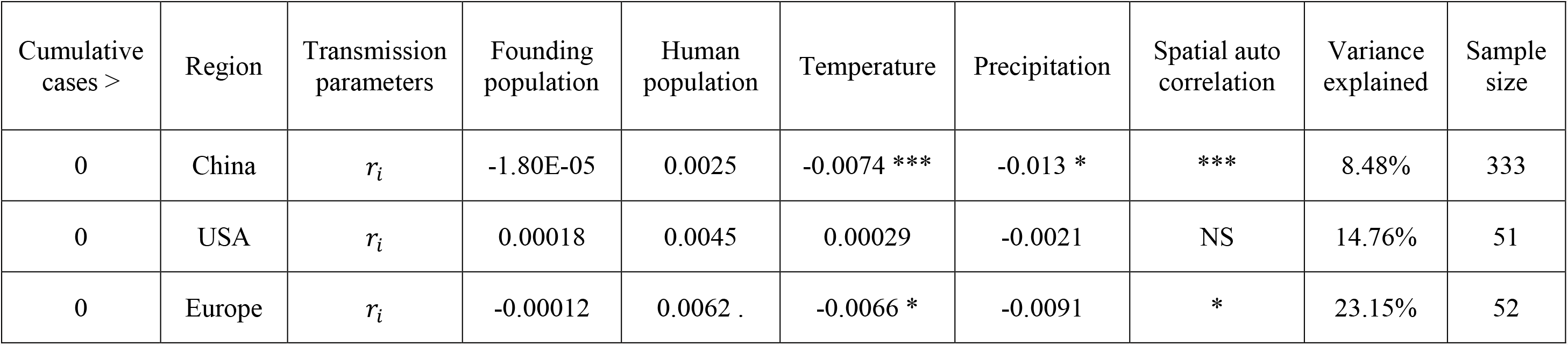
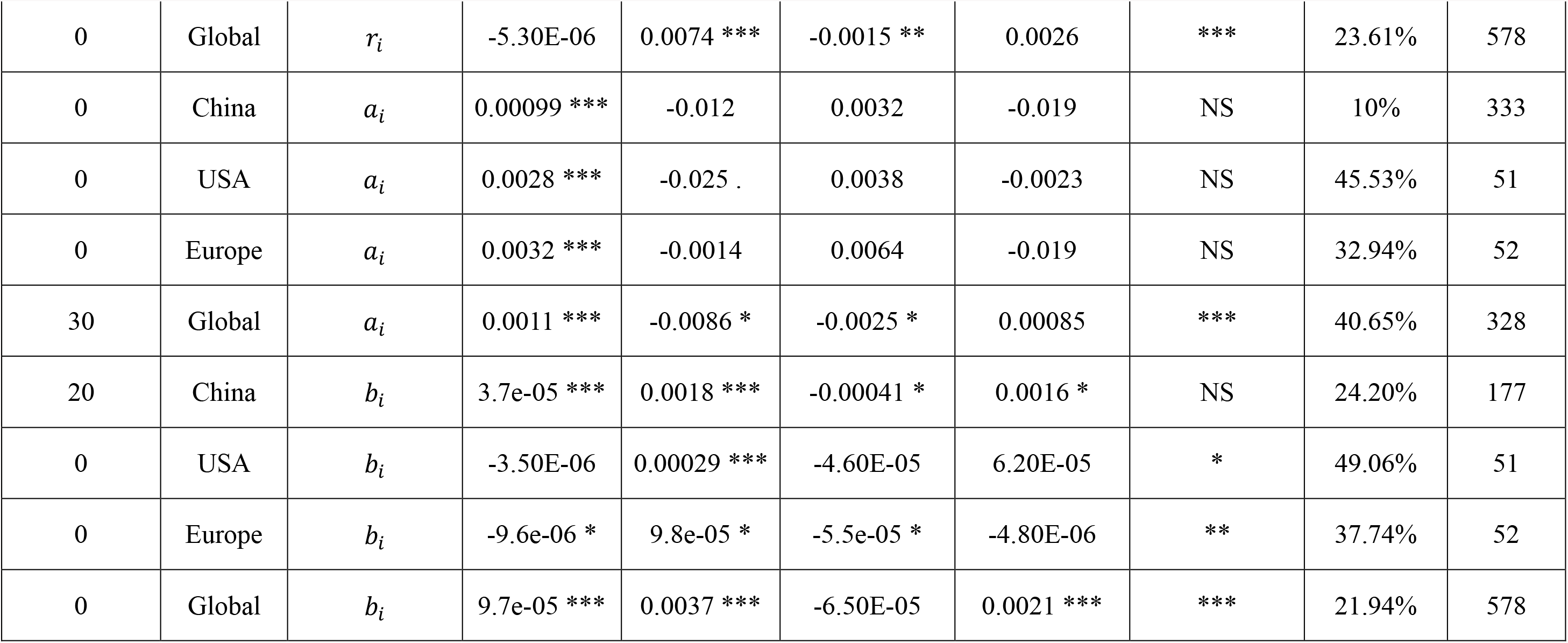
Significant associations of the average daily increase rate (*r*_i_) of cumulative cases, the maximum daily increase rate (*a*_i_), and the control efficiency (*b*_i_) with the founding population size of COVID-19 patients during the initial 7 days after the first reported patient(*F*_i_), human population size of a location (*H*_i_), climate factors (temperature,*T*_i_, and precipitation, *P*_i_), and spatial autocorrelation based on analyses using Equation 2 (linear model). + denotes the significant effects of spatial autocorrelation or temperature (*p* < 0.05), NS denotes non-significant effects. * *p* < 0.05, ** *p* < 0.01, *** *p* < 0.001. To obtain robust results, the linear model for each country or region was repeated four times with the cumulative cases >0, 10, 20, and 30, respectively (SI Appendix, Table S2). Models presented here were based on the observation that human or climate variables should have significant association with the average daily increase rate (*r*_i_), the maximum daily increase rate (*a*_i_) or the control efficiency (*b*_i_) in at least two repeated models from SI Appendix, Table S2.

Analysis using nonlinear GAM models (Fig. 2, SI Appendix, Table S1,S3) was very similar to those using the linear GAM model. Additionally, we found air temperature showed a negative and a weak dome-shaped association with the average daily increase rate peaked around –4.4°C (Fig. 2B), a positive association with the maximum daily increase rate of Europe (Fig. 2E), and a strong dome-shaped association with the maximum daily increase rate in the world which peaked around 6.3°C (Fig. 2F). A few previous studies also reported the nonlinear association of temperature with transmission severity of COVID-19 using incidence case (Bannister-Tyrrell et al., 2020; Bu et al., 2020; Chen et al., 2020; J. Wang et al., 2020) or growth rate (Ficetola & Rubolini, 2020; Notari, 2020), but the results were inconsistent. A few studies indicated that the optimal temperature for SARS-CoV-2 incidence of new cases was at 8.07 °C (Chen et al., 2020) or 8.72°C around the world (J. Wang et al., 2020), and 13∼19°C in China (Bu et al., 2020). The growth rates peaked at about 5°C in temperate regions in the Northern Hemisphere during the outbreak month, while they decreased in warmer and colder regions (Ficetola & Rubolini, 2020). Bannister-Tyrrell *et al*. (2020) found COVID-19 incidence had a belled-shaped association around 1°C (Bannister-Tyrrell et al., 2020). Notari (2020) reported that temperature had a negative association with early exponential growth with a weak peak at about 7.7±3.6°C (Notari, 2020). In our study, a strong dome-shaped association between the maximum daily increase rate of the world and air temperature that peaked around 6.3°C was identified, which was close to those (Ficetola & Rubolini, 2020; Notari, 2020) using similar measures, but the dome-shaped association was much stronger in our study. Our peak temperature was close to those using incidences of COVID-19 (Chen et al., 2020; J. Wang et al., 2020), however, after excluding the spatial autocorrelation and human factors, we did not find any significant association of the number of cumulative cases of COVID-19 with climate factors in China, USA, Europe and the world, although it had significant association with human factors (SI Appendix, Table S4).

**Figure 2.**
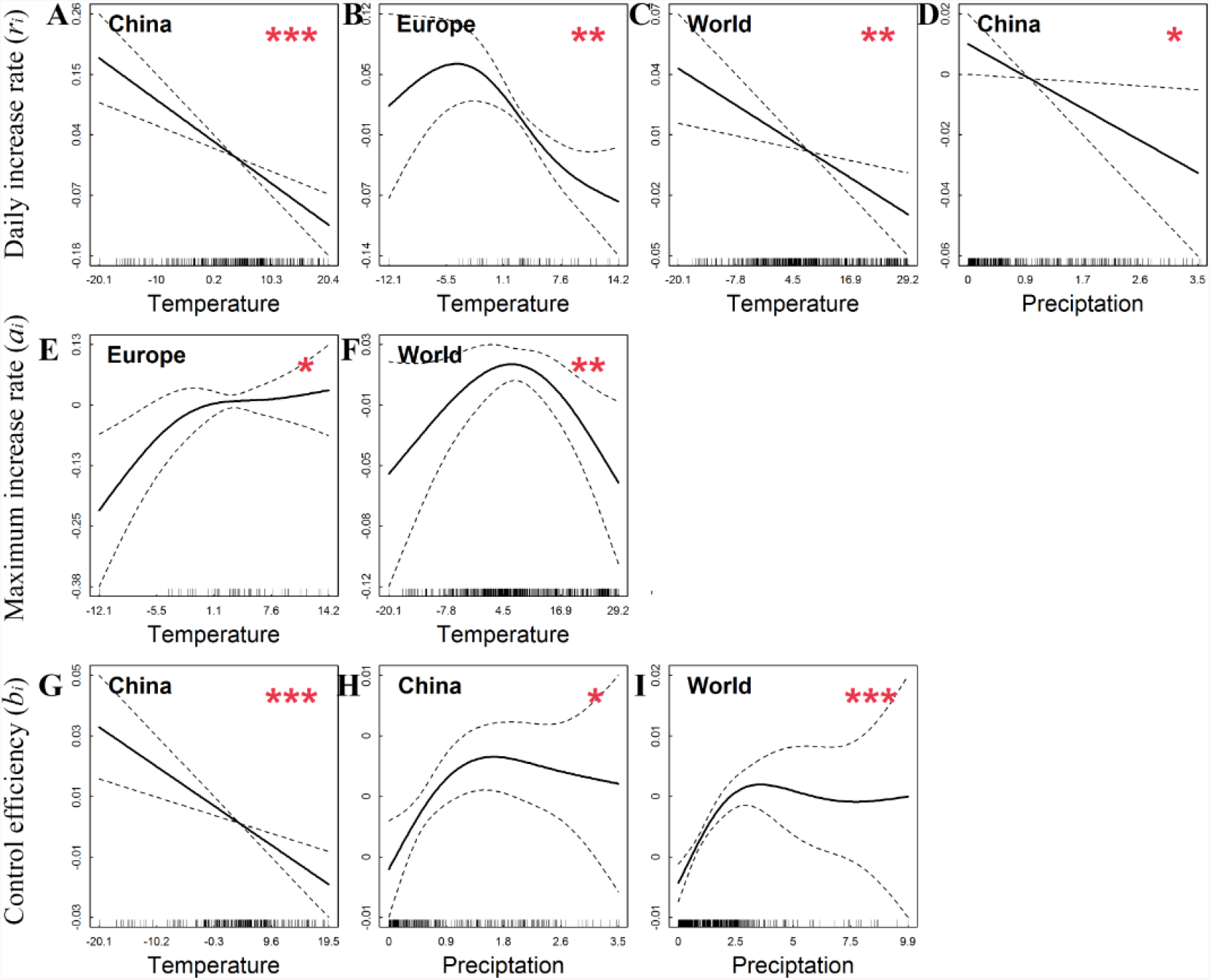
Significant partial effects of temperature and precipitation on daily increase rate (*r*_i_), maximum daily increase rate (*a*_i_), and control efficiency (*b*_i_)(Note: smaller bi means better control efficiency) based on results of the nonlinear models using Eq.3 after excluding the confoundingfactors of humans and spatial autocorrelation. A-C. Effects of temperature on A-C. Effects of temperatur *r*_i_ in China, Europe and the world.D. Effects of precipitation on *r*_i_ in China. E-F. Effects of temperature on in Europe and the World. G. Effects of temperature on *b*_i_ in China. H-I. Effects of precipitation on *b*_i_ in China and the World. Dashed lines are 95% nominal confidence bands. Solid lines are statistically significant (*p < 0.05, ** p < 0.05, *** p < 0.05). The country of region was shown in the top-left corner. The short and black vertical lines along x axis indicated the data distribution of temperature (°C) orprecipitation (mm/day).

The dome-shaped relation of organisms with environmental factors is reasonable based on the Law of Tolerance (Shelford, 1931). Chin *et al*. (2020) reported that the virus of COVID-19 was highly stable with only a 0.6-log unit reduction at 4°C in 14 days, with a 3-log unit reduction at 22°C after 7 days and no detection at 14 days, with a 3-log unit reduction at 37°C after 1 day and no virus detected afterwards (Chin et al., 2020). This observation supports our observation on the peak temperature of 6.3°C of the virus of COVID-19. Although the virus could be well preserved in cold conditions, the lower transmission ability under 6.3°C was likely caused by human behaviors. In cold conditions, people are not as active as in warm conditions, which did not favor the person to person transmission of COVID-19. Besides, in cold conditions, droplets can freeze, which prevent their spreading in the air.

The impacts of climate on control efficiency have never been assessed before. We found air temperature showed a significant and negative association with the control efficiency (*b*_i_) in China and Europe (Fig. 2G, Table 1). High precipitation showed a significant and positive association with the control efficiency (*b*_i_) in China and the world (Table 1). These results indicated that cold and wet climate decreased the control efficiency (Note: smaller *b*_i_ indicates the better control efficiency) on COVID-19. This is likely because cold and wet conditions did not favor human movement outside, thus the lockdown and social distancing measures may have worked better in warm and dry conditions rather than in cold and dry conditions. Another possible explanation could be that in cold conditions, people and facilities for disease control or prevention may not be easily mobilized. The efficiency of detection, disinfection or sanitation may be low in cold condition. Therefore, although the COVID-19 virus had an optimal temperature around 6.3°C, the poor control efficiency in cold conditions resulted in the general negative association of temperature with the average daily increase rate.

Our results suggest that climate may affect the transmission of COVID-19 directly on the viability of the virus, and indirectly through affecting human behavior as well as the control efficiency. Human immunity might be lower under cold conditions, which makes them more susceptible to the virus (Eccles, 2002; Kudo et al., 2019), however, human movement and contact may be low in cold conditions, which might reduce the risk of transmission. The virus appears unstable with high UV irradiation and high temperatures (Duan et al., 2003; Lowen & Steel, 2014; Tellier, 2009), but it may also be arrested in cold condition; thus extreme conditions would make it hard for the virus to survive or spread in cough droplets of infected patients or on the surfaces of contaminated goods. Besides, the control efficiency would be high in warm conditions but low in cold conditions, which might contribute to the observed association between climate and transmission ability.

### Impacts of human factors

The effects of human factors on the spread of COVID-19 and control efficiency have been widely modelled in previous studies (e.g. (Gilbert et al., 2020; Hellewell et al., 2020; Tian et al., 2020), but evidence using empirical data is still limited. In this study, we found the founding population size of early reported COVID-19 patients showed a consistent, significant and positive association with the maximum daily increase rate (*a*_i_) in China, USA, Europe and the world (Table 1). It had a positive association with the control efficiency (*b*_i_) in China and the world (but a negative association in Europe). These results suggested that human migration increased the transmission severity of COVID-19. A higher number of early reported COVID-19 cases stimulated the increase of control efforts in Europe, but not in China and the world. The population size of a location showed a consistent, significant and positive association with the average daily increase rate (*r*_i_) and the control efficiency (*b*_i_) of COVID-19 in the world (Table 1), indicating countries or regions with large susceptible population suffered high infection of COVID-19 but with a smaller control efficiency (Note: smaller *b*_i_ indicated the higher control efficiency). The spread of disease is very similar to the biological invasion defined by the Allee effect (Allee, Park, Emerson, Park, & Schmidt, 1959). The Allee effect suggested that founding population size was essential for the successful establishment of alien species (Liebhold & Bascompte, 2003). It has been widely supported in studies of biological invasions (Courchamp, Clutton-Brock, & Grenfell, 1999; Stephens & Sutherland, 1999), as well as by our observations.

### Implications for prevention

Our study suggested that both human and climate factors determined the spread of COVID-19 by altering the transmission ability of COVID-19 and its control efficiency. Temperature showed distinct effects on the early (without human intervention) and later stage (under human intervention) transmission of COVID-19. Based on our results in Fig. 2F, C, we projected the influences of seasonal change of temperature on the contagious risk of early transmission without or with little human intervention (Fig. 3A-D), and the later transmission under human intervention (SI Appendix, Fig. S1) in the world. Summer seasons would decrease of the early transmission risk of COVID-19 from low-latitude or low-altitude regions but increase the transmission risk in the high-latitude or high-altitude regions. The summer season would decrease the late transmission ability of the northern hemisphere but increase that of the southern hemisphere.

**Figure 3.**
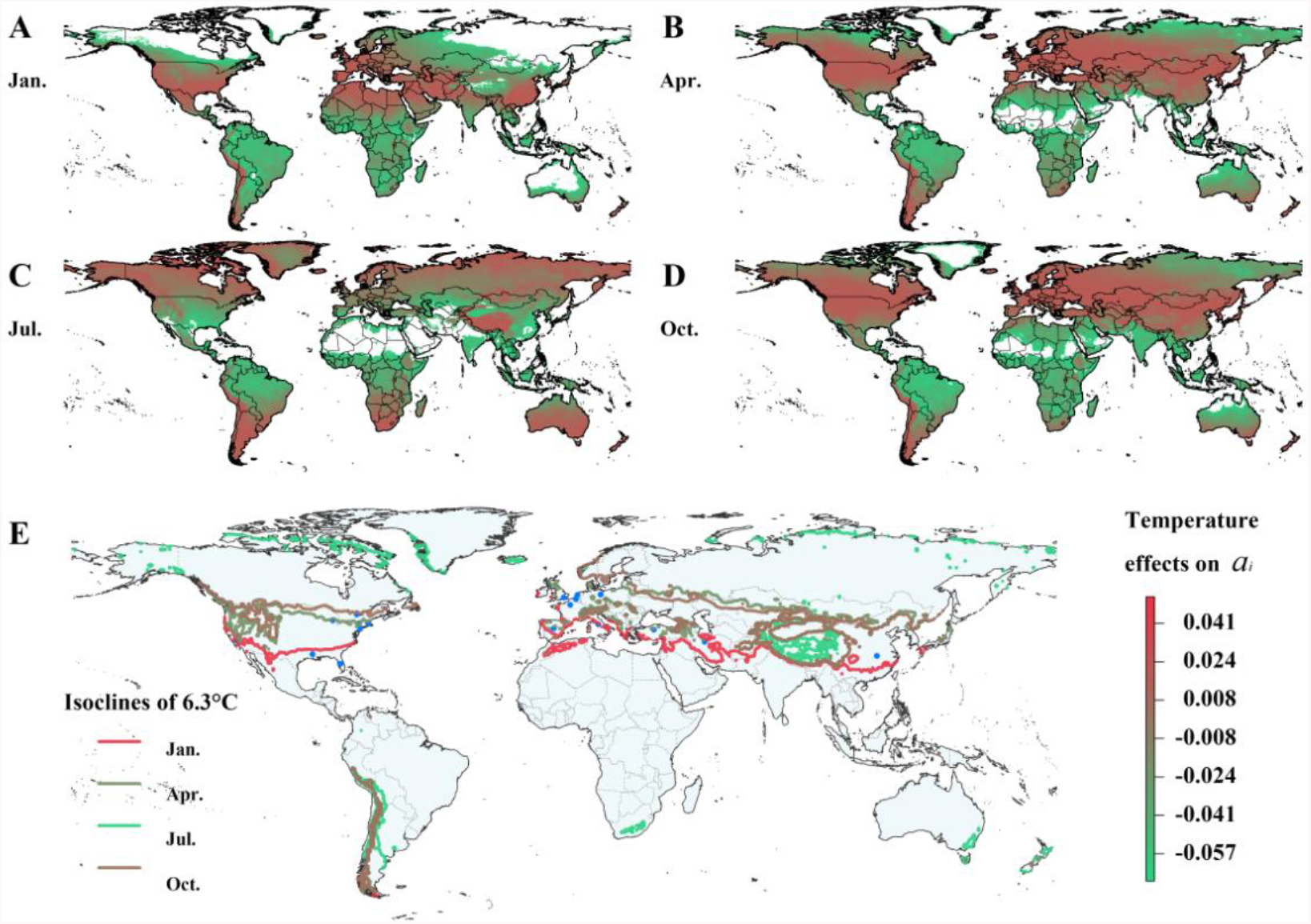
Seasonal projected partial effects of air temperature on the maximum daily increase rate (*a*_i_) without human intervention (A-D) and 6.3°C isoclines of COVID-19 in January, April, July and October (E). Colors (green to red: negative to positive) in Panel A-D indicate the temperature effects on maximum daily increase rate (*a*_i_) in different months, colors of isoclines in Panel E indicated different months.

We plotted the 6.3°C isocline of COVID-19 in January, April, July and October in Fig. 3E. It is notable that countries or regions with heavy infection rates of COVID-19 are mostly located within the isoclines of 6.3°C (the optimal temperature for COVID-19) from October to January. From winter, spring to summer, the 6.3°C isocline moves from subtropical to temperate, arctic zone; while from summer, autumn to winter, it back moves from arctic to temperate, subtropical zone (Fig. 3E). The seasonal movement of the 6.3°C isocline of COVID-19 along latitude or altitude increases the transmission risk of COVID-19 in different climate zones. Phylogenetic analysis suggested that the novel coronavirus was a close relation with the coronavirus of Middle East Respiratory Syndrome (MERS) and Severe Acute Respiratory Syndrome (SARS) (T.-n. O. J. F. E. I. Team & Li, 2020; Xu et al., 2020;Zhu et al., 2020). The whole genome of the COVID-19 virus has a high similarity(96%) with that of a coronavirus isolated from bats in Yunnan, China (Zhu et al.,2020). However, the natural hosts carrying the virus of COVID-19 and their habitats remain unclear. The area with the 6.3°C isocline between October and January shouldbe investigated as a priority in the search for the natural hosts of COVID-19, their habitats and migration routes.

Our study indicated that both human and climate factors played a significant role in the spread of COVID-19. It is not wise to rely upon climate factors to control this dangerous virus. Human intervention, such as lockdown and travel restrictions at the epicenter, as well as identification and isolation of infected patients or people with close contact, have been demonstrated to be successful in preventing the spread of COVID-19 (Tian et al., 2020). Thus, human intervention is essential to contain the rapid expansion of COVID-19 around the world. More efforts and collaboration are urgently needed in containing the spread of COVID-19 around the world.

## METHODS

### Epidemic data

We obtained data of cumulative cases of COVID-19 in cities and prefectures in China from 1 January to 11 March from daily reports or announcements by each provincial or prefectural health commission (making up 99.23%), the World Health Organization (making up 0.61%), and news from official media such as the CCTV news channel (making up 0.10%), and announcements by local governments (0.06%). Data consisted of the following information: reference, date, province, prefecture, coordinates, and cumulative case. The latitude and longitude coordinates of geographical locations were assigned by their capital site using a Baidu map (lbsyun.baidu.com). The data covered each prefecture of 27 provinces and autonomous regions, each district or county of 4 central municipal cities (i.e. Beijing, Tianjin, Shanghai, and Chongqing), 2 special administrative regions (Hong Kong and Macau SAR) and Taiwan. Cumulative cases in China after 11 March were not used because the daily increase rate after 11 March was smaller than 1%, indicating the approximate end of an epidemic. This data selection (the same as the other time series below) avoided biased estimation on the average, maximum daily increase rate and control efficiency with excessive data when the epidemic was close to an end.

We obtained data of cumulative COVID-19 cases of other countries, territories or regions from 20 January to 4 April from COVID–2019 situation reports by the World Health Organization on their website (https://www.who.int/emergencies/diseases/novel-coronavirus-2019/situation-reports/). We obtained the cumulative cases of COVID-19 of 50 states and the District of Colombia in the United States from 22 January to 4 April from Systems Science and Engineering (CSSE) at Johns Hopkins University (JHU) (https://www.arcgis.com/apps/opsdashboard/index.html#/bda7594740fd40299423467b48e9ecf6) (Dong, Du, & Gardner). Data consisted of date, country/territory/region, coordinates, cumulative cases and transmission category. The coordinates were assigned by referring to the capital city of each country or region using Google maps (https://www.google.com/maps/) For China, Japan, South Korea and Thailand, there are some missing cumulative case values (making up 0.71%); we assigned these missing values with those of the previous day. The model analysis was conducted separately for China, the USA, Europe, and the rest of the world, which represent the incidence of COVID-19 of three large epicenters and the world (Fig. 1, Table 1, SI Appendix, Table S1–3). Similarly, cumulative case data was not used for countries or regions when the daily increase rate was smaller than 1%. Cumulative cases were normalized by (average value-minimum value)/range of the value for easily demonstrating the growth patterns of different locations in Fig. 1, whereas the original data of cumulative cases was used for modeling analysis. It is notable that our data had various spatial resolution from prefecture to state or countries. However, spatial resolution was relatively comparable within China, USA and Europe.

### Anthropogenic and climate proxy data

The human population size (*H*) of a city or prefecture was obtained from China Population & Employment Statistics Yearbook 2018 complied by the Population and Employment Statistics Division, National Bureau of Statistics of China. The human population size of countries outside China was obtained from the World Bank (https://data.worldbank.org/indicator/SP.POP.TOTL). Gridded human population density was obtained from the Socioeconomic Data and Applications Center (SEDAC) (https://sedac.ciesin.columbia.edu/data/set/gpw-v4-admin-unit-center-pointspopulation-estimates-rev11) (Center for International Earth Science Information Network, 2018). Prefectural GDP in China was obtained from the provincial and national bureau of statistics in China, GDP of each state and District of Columbia in the USA was obtained from the U.S. Bureau of Economic Analysis (https://www.bea.gov/data/gdp/gdp-state) and the GDP of each country or region was obtained from World Bank (https://data.worldbank.org/indicator/NY.GDP.PCAP.CD). Human population size was log transformed (with base = e) to make the data normally distributed.

The daily average air temperature and 20:00–20:00 cumulative precipitation (during 2010–2019) from Chinese surface meteorological stations in China were obtained from the dataset of daily surface observation values (http://data.cma.cn/data/cdcdetail/dataCode/SURF_CLI_CHN_MUL_DAY_V3.0.html) (Ren et al., 2020), which was derived from daily report by 699 meteorological stations. The temperature and precipitation of each location (a sum of 333 locations in this study) in China was assigned by the temperature and precipitation of the nearest meteorological station. The average daily air temperature (*T*) and average daily precipitation (*p*) of an epidemic of a location was calculated by assigning the date of incidence of COVID-19 to the date of corresponding month of the historical data during 2010–2019.

We obtained monthly average temperature and precipitation (during 1970–2000) with a spatial resolution of 5 minutes from Worldclim 2 as climate proxy of countries except for China (Fick & Hijmans, 2017), the temperature and precipitation of each country was assigned by the temperature and precipitation of its capital city. Monthly precipitation was transferred to daily precipitation by dividing by 30 days in further analysis.

### Estimation of transmission ability and control efficiency of COVID-19

We used a logistic model to estimate the transmission parameters of COVID-19 by following our previous study on SARS (Zhang, Sheng, Ma, & Li, 2004). The logistic model is widely used for studying population dynamics of animals (Krebs, 2014). The number of cumulative cases of COVID-19 can be well fitted as follow:

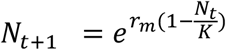

*N*_t_ was the number of cumulative cases at day *t*, *K* was the maximum cumulative cases of COVID-19 patients. The daily increase rate (*r*_t_) of the number of cumulative cases of patients was defined as follow:

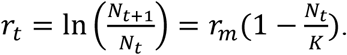

Thus, the daily increase rate should be negatively associated with the number of cumulative cases of patients under human intervention:

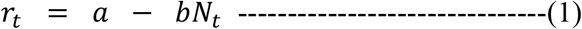

Here, *a*,*b* are constants, and all > 0. *a* represents the maximum daily increase rate (*r*_m_) without human intervention, *b* represents the control efficiency under human intervention.

Because the mean incubation period of COVID-19 patients was estimated to be 5.2 days (Li et al., 2020), and the incubation time of SARS was about 5 or 6.4 days (Lipsitch et al., 2003), we defined the number of cumulative cases of COVID-19 of a location in the first week as the founding population size (*F*) of early reported COVID-19 reported cases. We removed data of cumulative cases when the daily increase rate was less than 1% of the cumulative cases, intending to remove excessive data after the approximate end of an epidemic. We estimated the transmission parameters by using equation 1 and data of cumulative cases of an epidemic of a location covering the period from the 7^th^ day to the date of last observation or when the daily increase rate was less than 1% of the cumulative cases. Five locations (making up 0.86% of the data) with a < 0 (indicating minus maximum daily increase rate due to small sample size, no biological meaning) was removed in the analysis. A total of 578 time series of cumulative cases of COVID-19 from China (n = 333, 1 January −11 March), USA (n = 51, 22 January− 4 April), Europe (n = 52, 25 January− 4^th^ April) and the rest of world (n = 142, 20 January−;4 April) was constructed for estimating the transmission parameters (Fig. 1).

### Statistical Analysis

The process of a virus invading a new place is likely similar to the biological invasion of alien species. The founding population is very essential for the successful invasion as defined by the Allee effect (Allee et al., 1959). Besides, the population size and climate factors may also play a significant role in affecting the spread of COVID-19 both directly and indirectly. Thus, we assumed that the transmission parameters (*r*_i_, *a*_i_,*b*_i_) should be determined by the founding population size (*F*_i_), human population size of a location (*H*_i_), air temperature (*T*_i_) and precipitation (*P*_i_) in an *i*^th^ location. *r*_i_, *a*_i_ and *b*_i_ represent the average daily increase rate of cumulative cases, maximum daily increase rate, and control efficiency of COVID-19, respectively. *r*_i_ represents the transmission ability of COVID-19 under human intervention. *a*_i_ represents the maximum transmission ability (*r*_m_) without human intervention. *b*_i_ represents the control efficiency under human intervention. Because GDP per capita, population density had strong correlation with the population size (*r* = 0.67 for China, *r* = 0.84 for USA), thus, we only used population size for analysis.

GAMs were used to model the effects of the founding population size (*F*_i_), human population size (*F*_i_), and climate factors (air temperature *T*_i_ and precipitation *P*_i_) on the average daily increase rate (*r*_i_), maximum daily increase rate (*a*_i_), and control efficiency (*b*_i_) in the *i*^th^ location by following (Wood, 2011). A Gaussian GAMs was firstly fitted by using a linear regression formula:

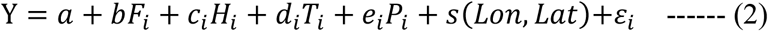

Here, *Y* represents the three transmission parameters (*r*_i_,*a*_i_,*b*_i_) separately. To examine the potential nonlinear effect of climate factors, we fitted the data using the following model:

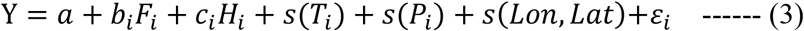

Here, *s*(*T*_i_), *s*(*P*_i_)and *s*(*Lon,Lat*) were 2D smooth function (with *k* value, dimension of the basis = 4) for removing the effects of spatial autocorrelation. ⁵_𝑖_ was uncorrelated random errors of zero mean and finite variance. To obtained model robustness as to the effects of human and climate factors on the transmission of COVID-19, we conducted the modeling analysis by using the cumulative cases > 0, 10, 20 and 30 infected cases of COVID-19 (SI Appendix, Table S2, S3). Only significant effects of a factor on a transmission parameter detected in two modeling analyses were selected for making conclusions and discussions (Table 1, SI Appendix, Table S1). Using equation (2), 3), we also analyzed the association of the number of cumulative cases with human and climate factors (SI Appendix, Table S4).

Pearson’s correlation analysis was introduced to detect significant correlations among variables (SI Appendix, Fig. S2–S5). Loess regression was introduced to show the changing trend of cumulative cases of COVID-19 in Fig. 1E-H. The relation of average or daily increase rate and control efficiency with climate factors were shown in Fig. S6–7 (not partial relation). For variables with strong and significant correlations (*r* < –0.6 or *r* > 0.6; *p* < 0.05) with the other variables, only one variable with the largest correlation coefficient to transmission parameters was selected to avoid the potential collinearity effect in model analysis. GAM was carried out using the mgcv library (v.1.8–15) (Wood, 2011) in R (v. 3.6.1). Associations of the average daily increase rate or the number of cumulative cases with environmental variables of each city or prefecture were analyzed by using raster (v.2.9–22) and rgdal (v.1.4–4) libraries in R (v. 3.6.1) (Bivand, Keitt, & Rowlingson, 2016). Correlation analysis, linear regression, and loess regression was performed with the stats library (v. 3.6.1) in R (v. 3.6.1) (R. C. Team,2019).

## Data Availability

The data will be released upon the official publication of this article.

## ACKNOWLEDGEMENTS

This study was supported by the project of studying transmissions of COVID-19 and risk assessments based on big data analysis supported by the Institute of Zoology, Chinese Academy of Sciences. **Author contributions:** ZZ designed the study, XW and CC collected the data. XW did the analysis. All authors contributed intellectually to the manuscript. The authors declare no conflict of interest. **Ethical statement:** All data related to COVID-19 patients in this this study is obtained from the official website of China’s National Health Commission, WHO and the other published databases. All data in this study do not contain private or unpublished information of patients.

## Supplementary Information

Correlation between the transmission severity and human/climate factors

Using Pearson’s correlation analysis on data from 1 January to 11 March in China, we found the average daily increase rate was negative correlated with temperature (*r* =–0.11, *p* < 0.05), the maximum daily increase rate was positive correlated with founding population size during the initial 7 days after the first patient (*r* = 0.28, *p* < 0.001), the control efficiency was positive correlated with founding population size during the initial 7 days after the first patient (*r* = 0.21, *p* < 0.001), human population (*r* = 0.19, *p* < 0.001), and precipitation (*r* = 0.24, *p* < 0.001) (Fig. S2).

**Figure S1.**
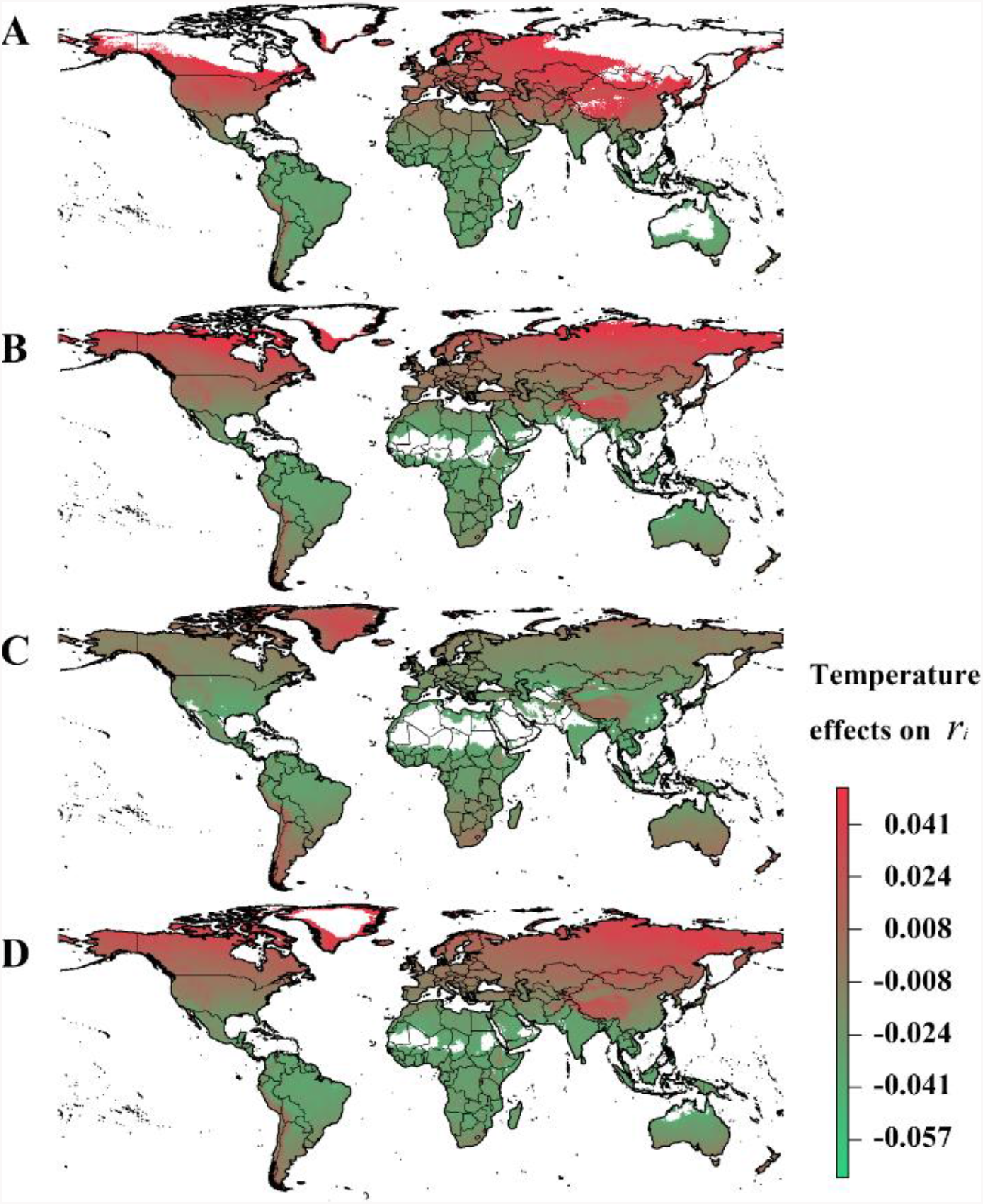
Seasonal projected partial effects of air temperature on the average daily increase rate (*r*_i_) under human intervention in January (A), April (B), July (C) and October (D). Colors (green to red: negative to positive) indicate the temperature effects on average daily increase rate(*r*_i_).

**Figure S2.**
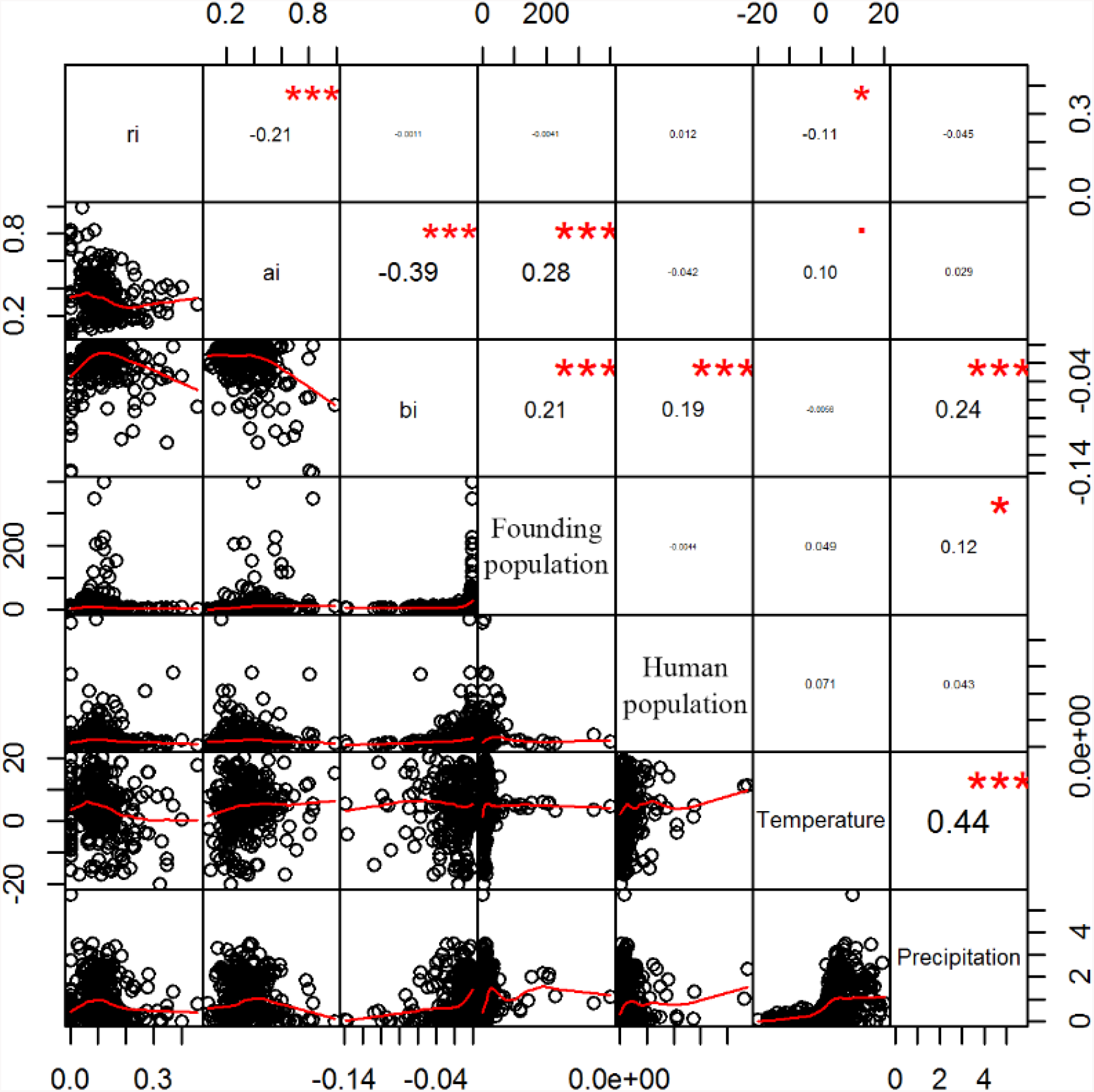
Correlation of the average daily increase rate(*r*_i_) of cumulative cases, maximum daily increase rate (*a*_i_), and control efficiency (*b*_i_) of COVID-19 with founding population size during the initial 7 days after the first patient, human population, and climate factors (temperature and precipitation) in China.

Using Pearson’s correlation analysis on data from 20 January to 4 April in the USA, we found the maximum daily increase rate was positive correlated with founding population size during the initial 7 days after the first patient (*r* = 0.63, *p* < 0.001), the control efficiency was positive correlated with human population (*r* = 0.41, *p* < 0.01), and precipitation (*r* = 0.40, *p* < 0.01) (Fig. S3).

**Figure S3.**
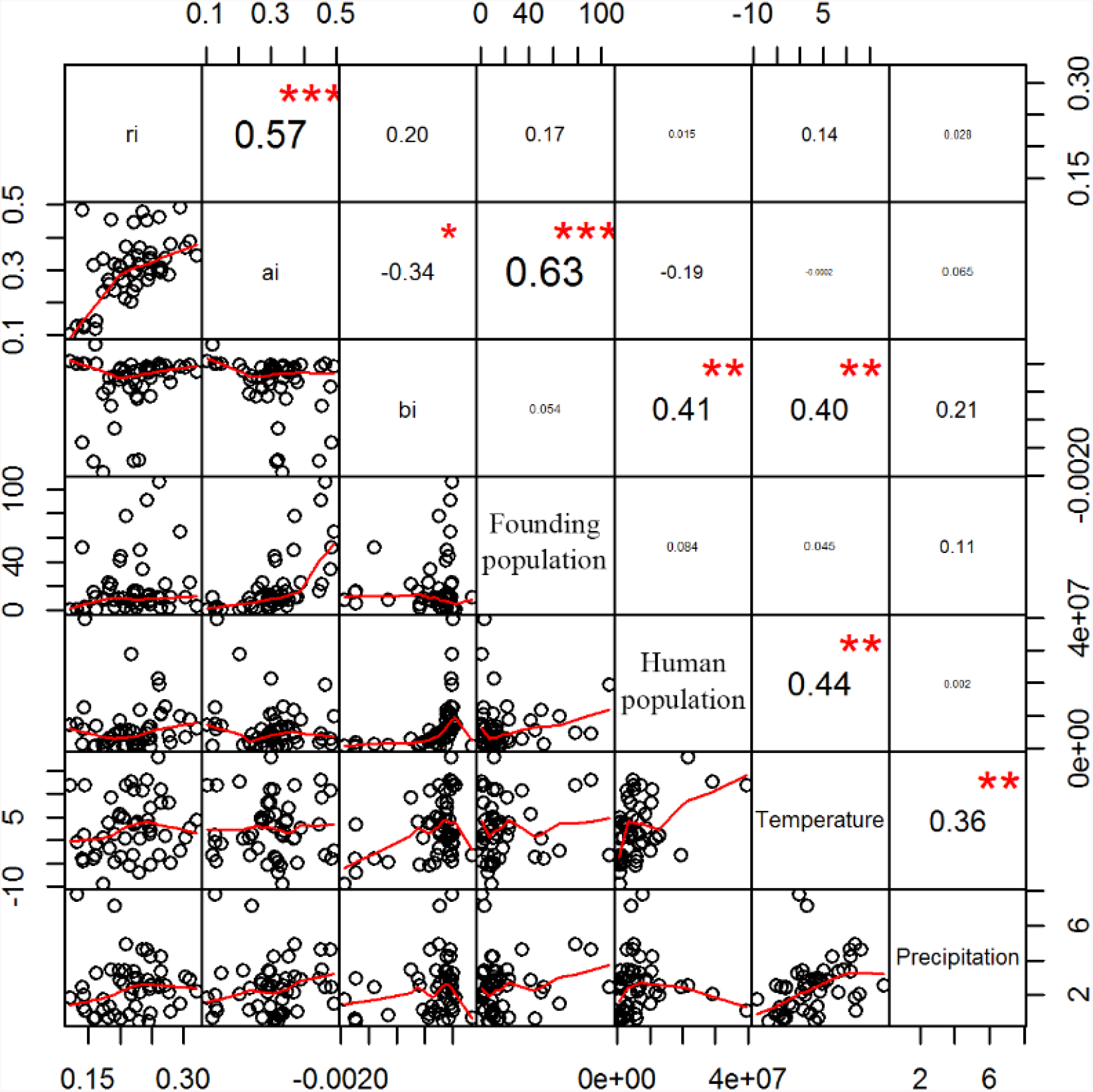
Correlation of the average daily increase rate (*r*_i_) of cumulative cases, maximum daily increase rate (*a*_i_), and control efficiency (*b*_i_) of COVID-19 with founding population size during the initial 7 days after the first patient, human population, and climate factors (temperature and precipitation) in the USA.

Using Pearson’s correlation analysis on data from 20 January to 4 April in Europe, we found the maximum daily increase rate was positive correlated with founding population size during the initial 7 days after the first patient (*r* = 0.49, *p* < 0.001), the control efficiency was positive correlated with founding population size during the initial 7 days after the first patient (*r* = –0.33, *p* < 0.05) (Fig. S4).

**Figure S4.**
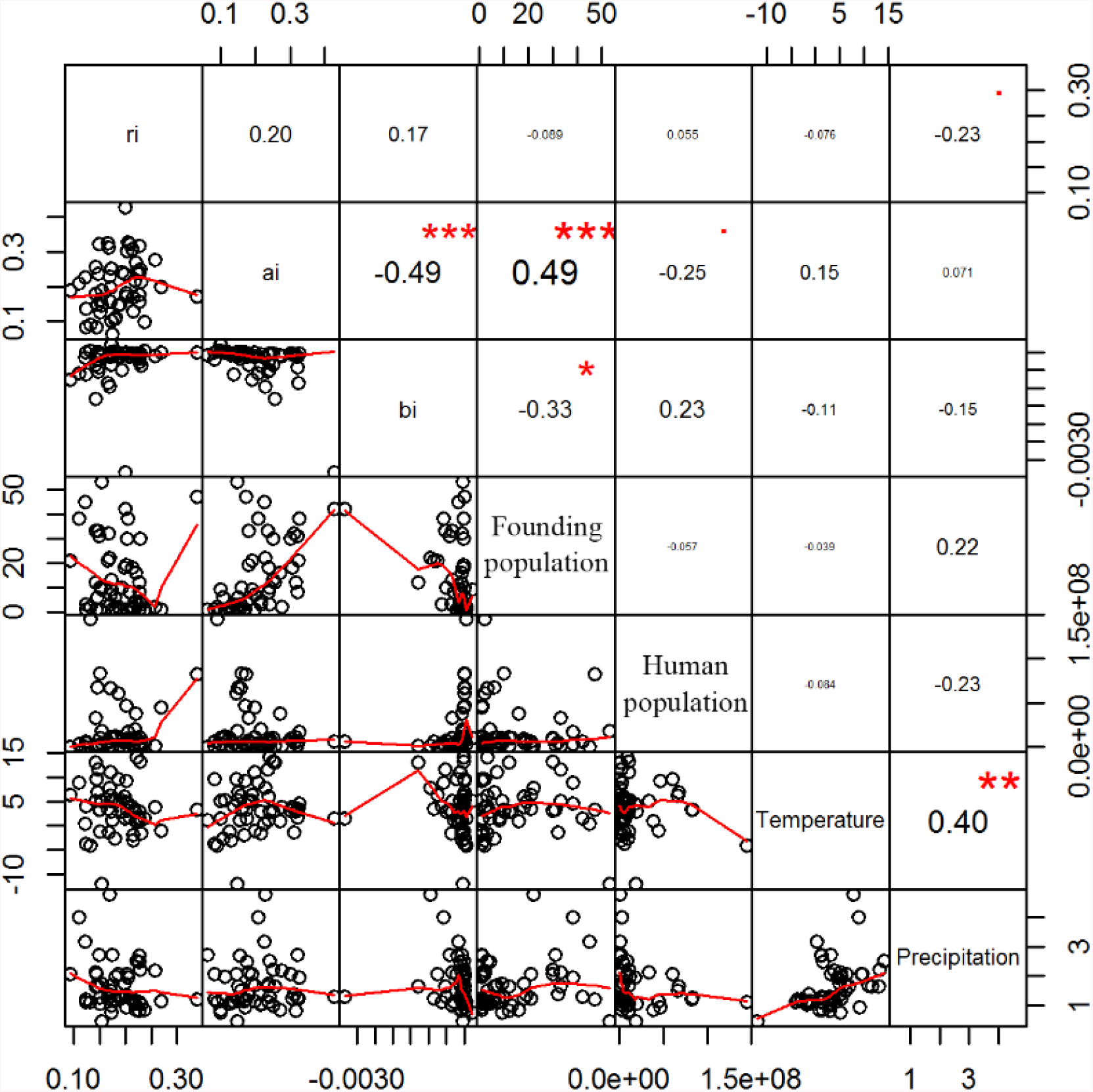
Correlation the average daily increase rate (*r*_i_)of cumulative cases, maximum daily increase rate (*a*_i_), control efficiency (*b*_i_) of COVID-19 with founding population size during the initial 7 days after the first patient, human population, and climate factors (temperature and precipitation) in Europe.

Using Pearson’s correlation analysis on data from 1 January to 4 April for the world data, we found the average daily increase rate was positive correlated with precipitation (*r* = 0.11, *p* < 0.01), maximum daily increase rate was positive correlated with founding population size during the initial 7 days after the first patient (*r* = 0.30, *p* < 0.001), but negative correlated with human population (*r* = –0.14, *p* < 0.01), temperature (*r* = –0.22, *p* < 0.001), and precipitation (*r* = –0.12, *p* < 0. 01), the control efficiency was positive correlated with founding population size during the initial 7 days after the first patient (*r* = 0.15, *p* < 0.001), temperature (*r* = 0.10, *p* < 0.05), and precipitation (*r* = 0.19, *p* 0.001) (Fig. S5).

**Figure S5.**
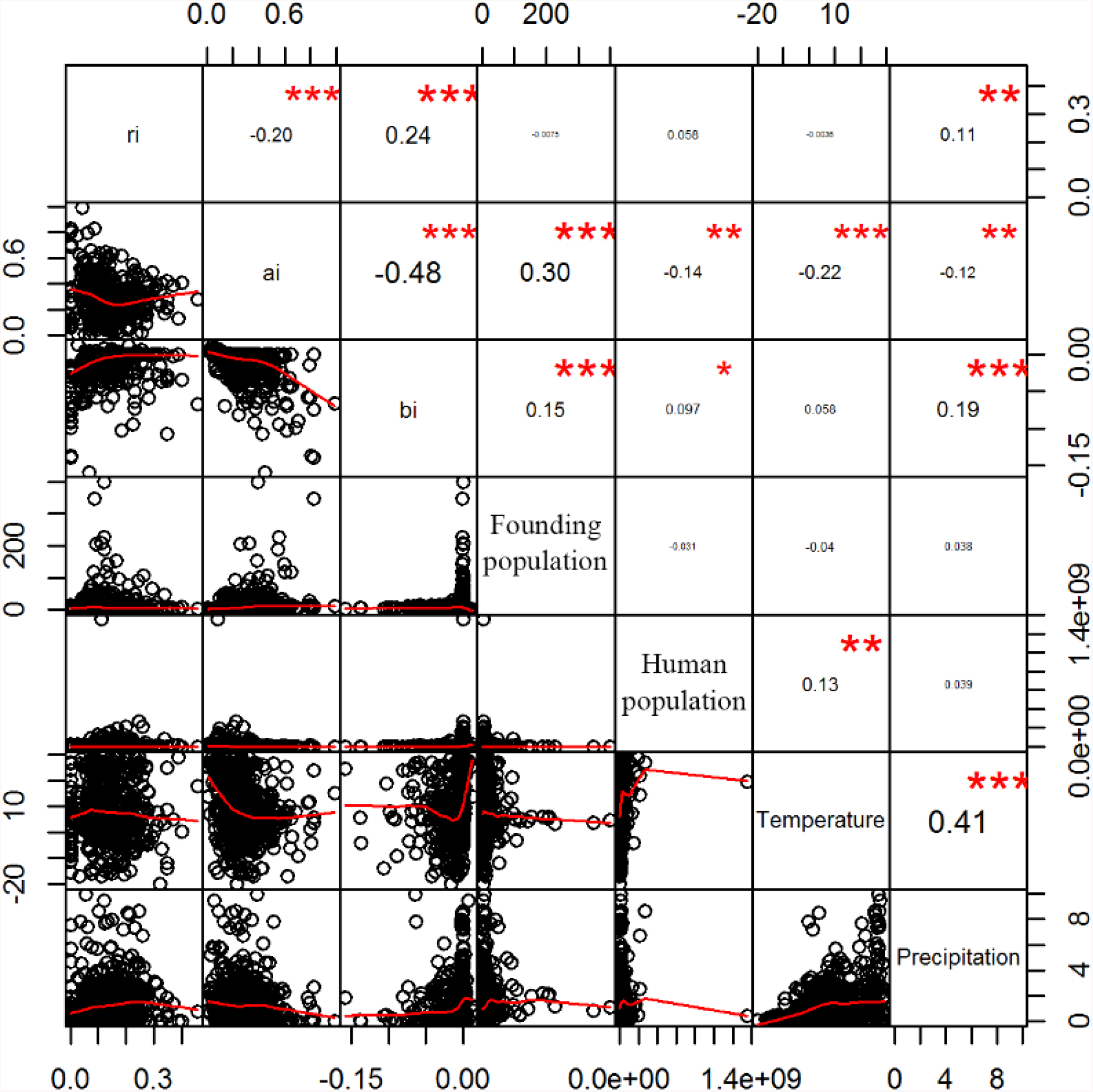
Correlation of the average daily increase rate (*r*_i_) of cumulative cases, maximum daily increase rate (*a*_i_), control efficiency (*b*_i_) of COVID-19 with the founding population size during the initial 7 days after the first patient, human population, and climate factors (temperature and precipitation) in the world.

**Figure S6.**
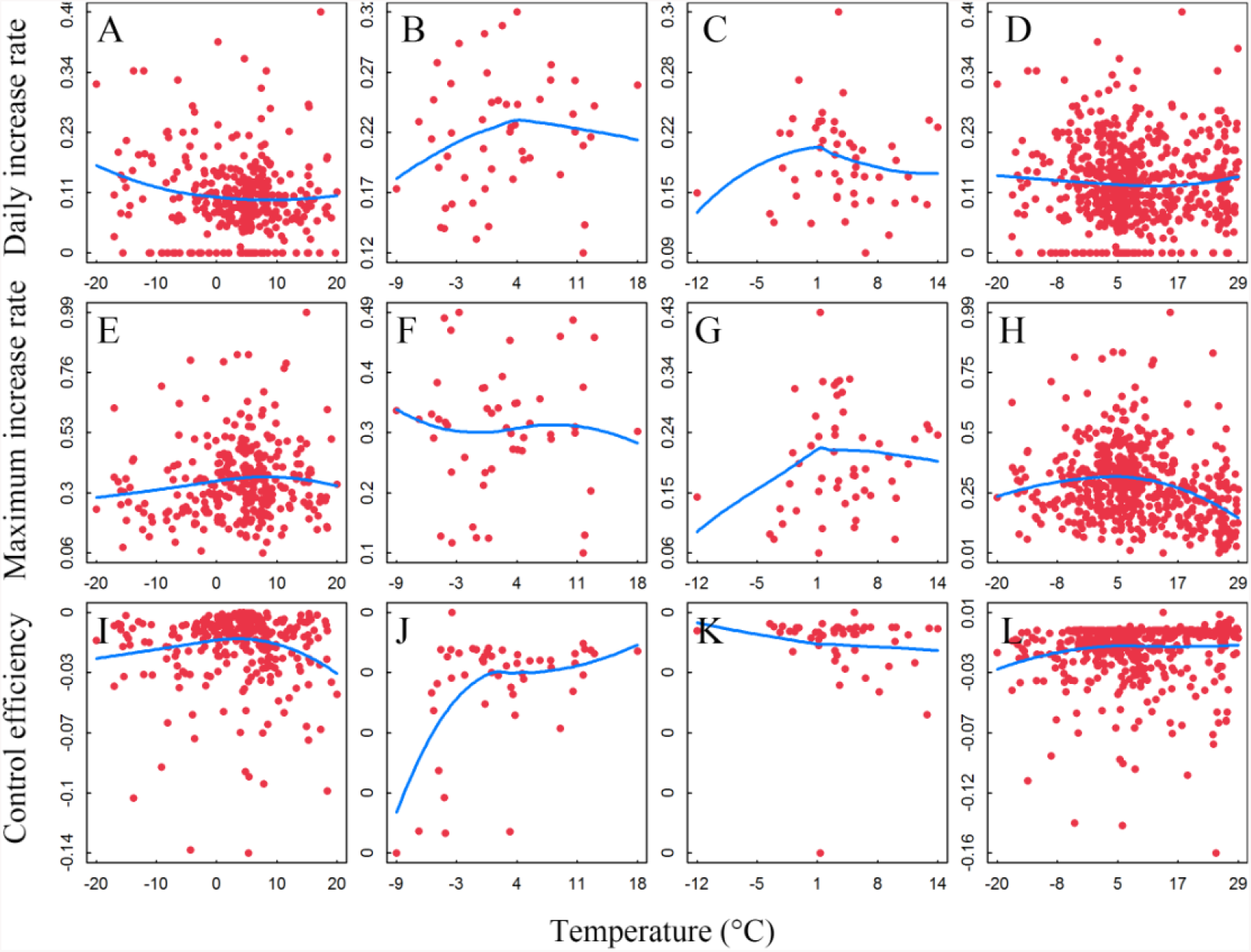
Relationship of temperature with the average daily increase rate (*r*_i_), maximum daily increase rate (*a*_i_), control efficiency (*b*_i_) of COVID-19 in China (A, E, I), USA (B, F, J), Europe (C, G, K), and the world (D, H, L). The blue lines indicate the loess regression with span = 1.

**Figure S7.**
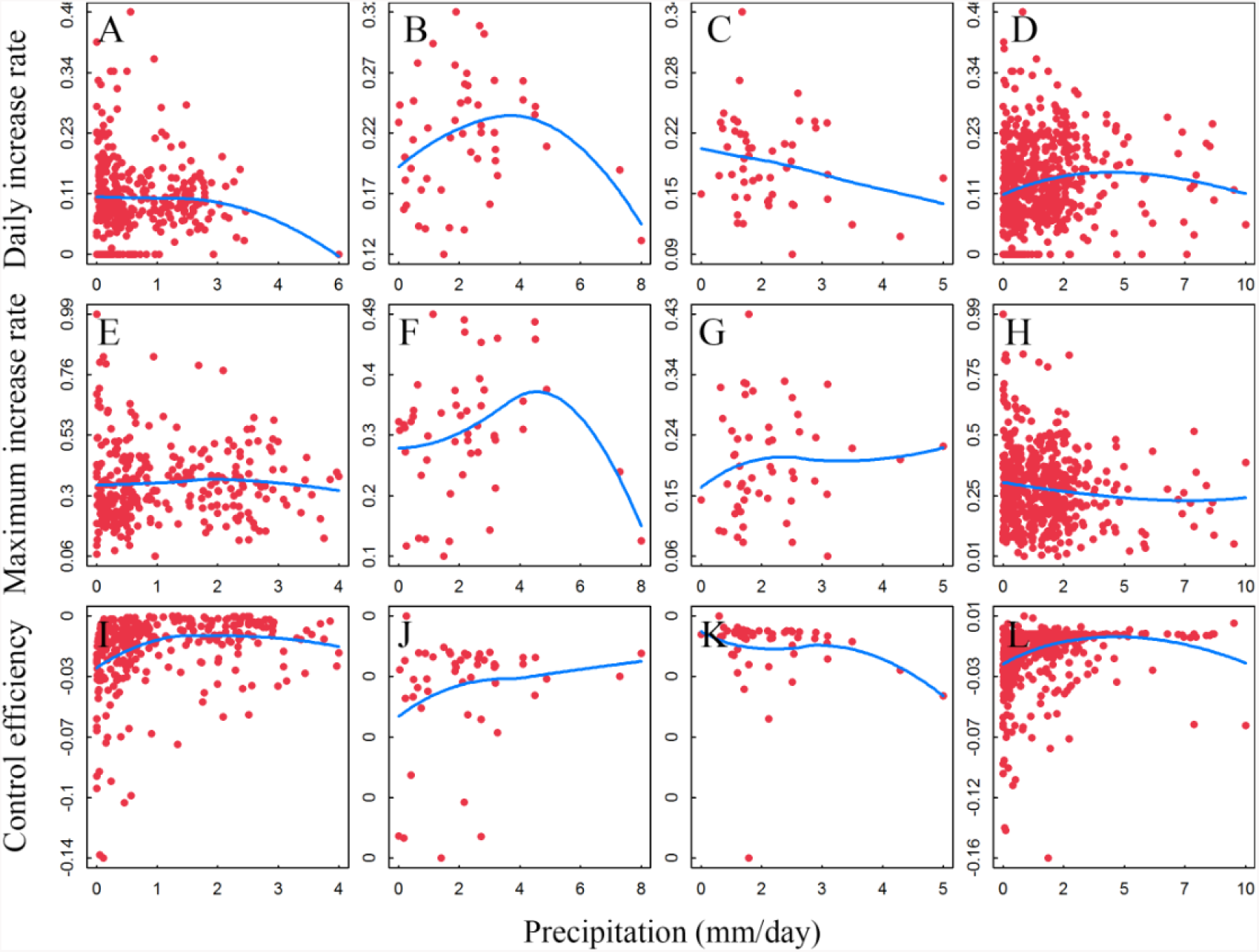
Relationship of precipitation with the average daily increase rate (*r*_i_), maximum daily increase rate (*a*_i_), control efficiency (*b*_i_) of COVID-19 in China (A, E, I), USA (B, F, J), Europe (C, G, K), and the world (D, H, L). The blue lines indicate loess regression with span = 1.

**Table S1:**
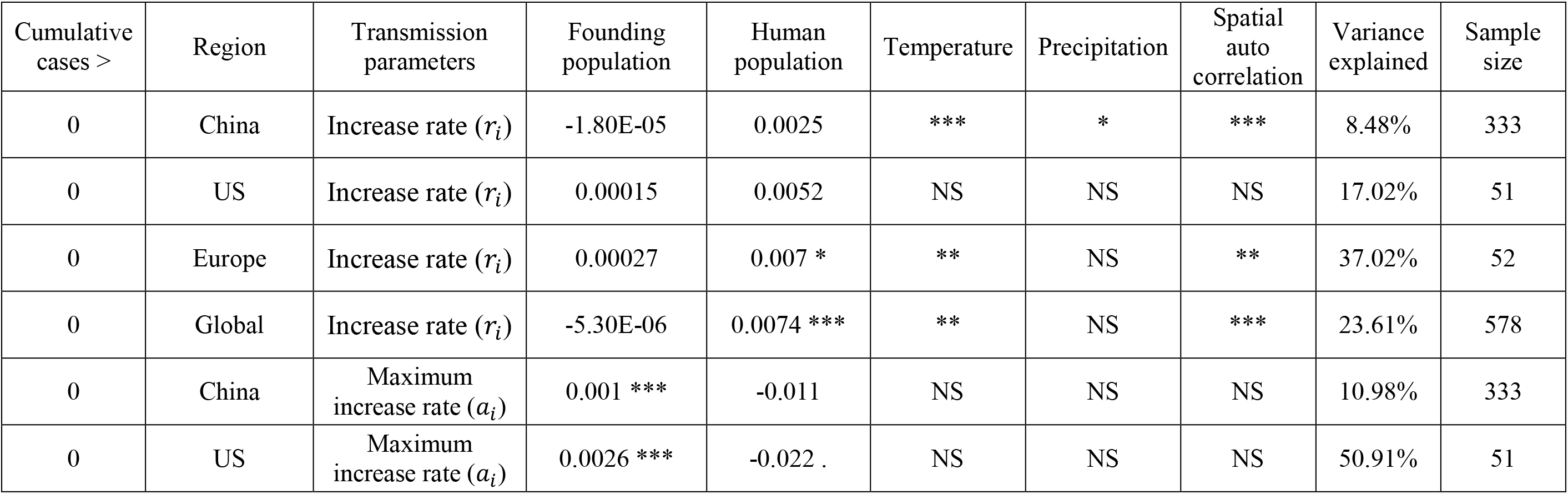
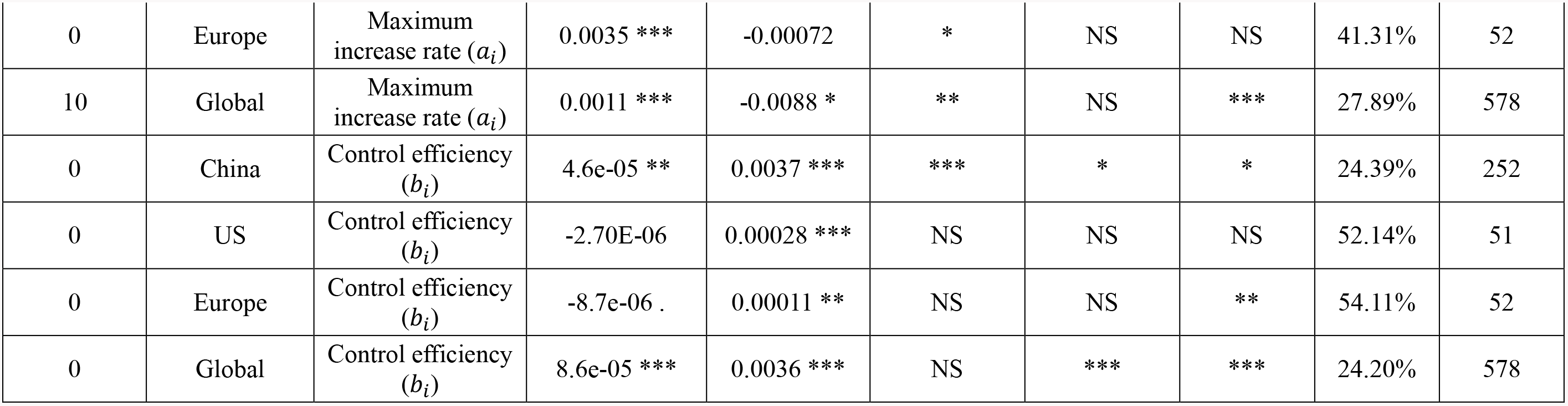
Significant associations of average daily increase rate (*r*_i_) of cumulative cases, maximum daily increase rate (*a*_i_), control efficiency (*b*_i_) of COVID-19 with founding population size during the initial 7 days after the first patient (*F*_i_), human population, climate factors (temperature and precipitation), and spatial autocorrelation. Bold values indicated the coefficients represent the significant effects(*p* < 0.05). + denotes the significant effects of spatial autocorrelation or temperature (*p* < 0.05), ns denotes non-significant effects. NA denotes not available (* *p* < 0.05, ** *p* < 0.01, *** *p* < 0.001). The significant association was extracted from Table S3. Models presented here were based on the observation that human or climate variables should have significant association with the average daily increase rate (*r*_i_), the maximum daily increase rate (*a*_i_) or the control efficiency (*b*_i_) in at least two repeated models from Table S3.

**Table S2:**
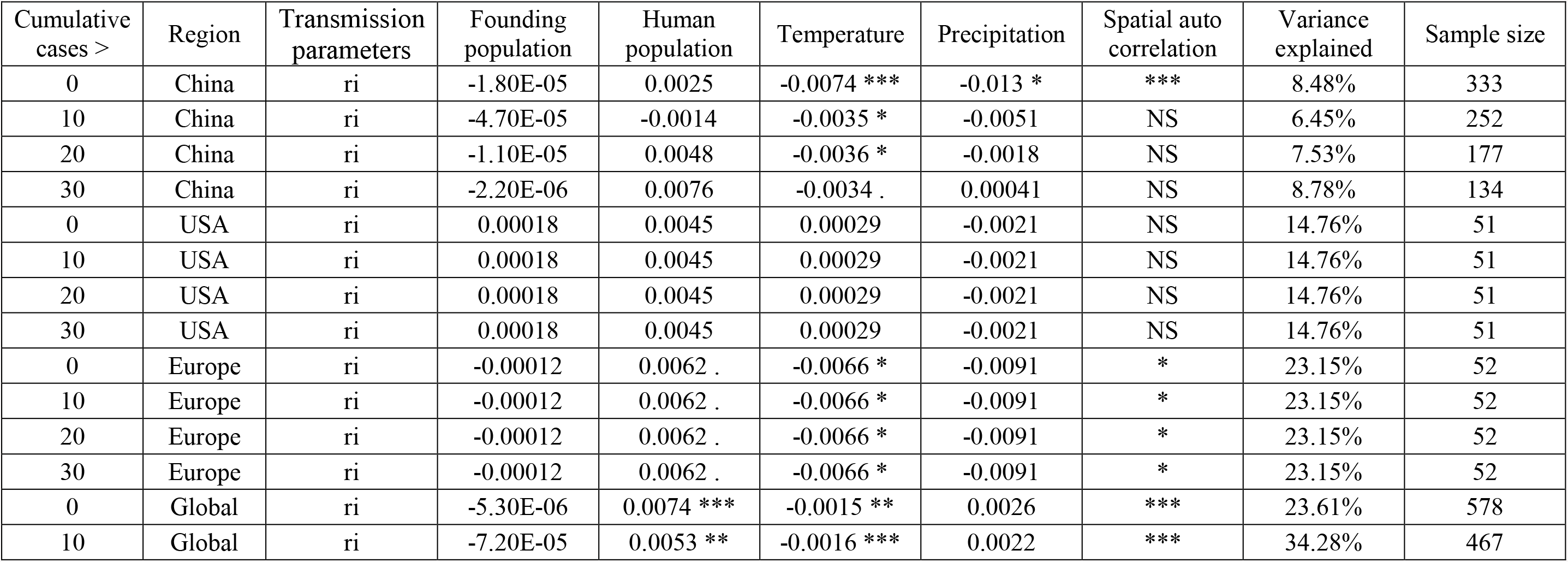
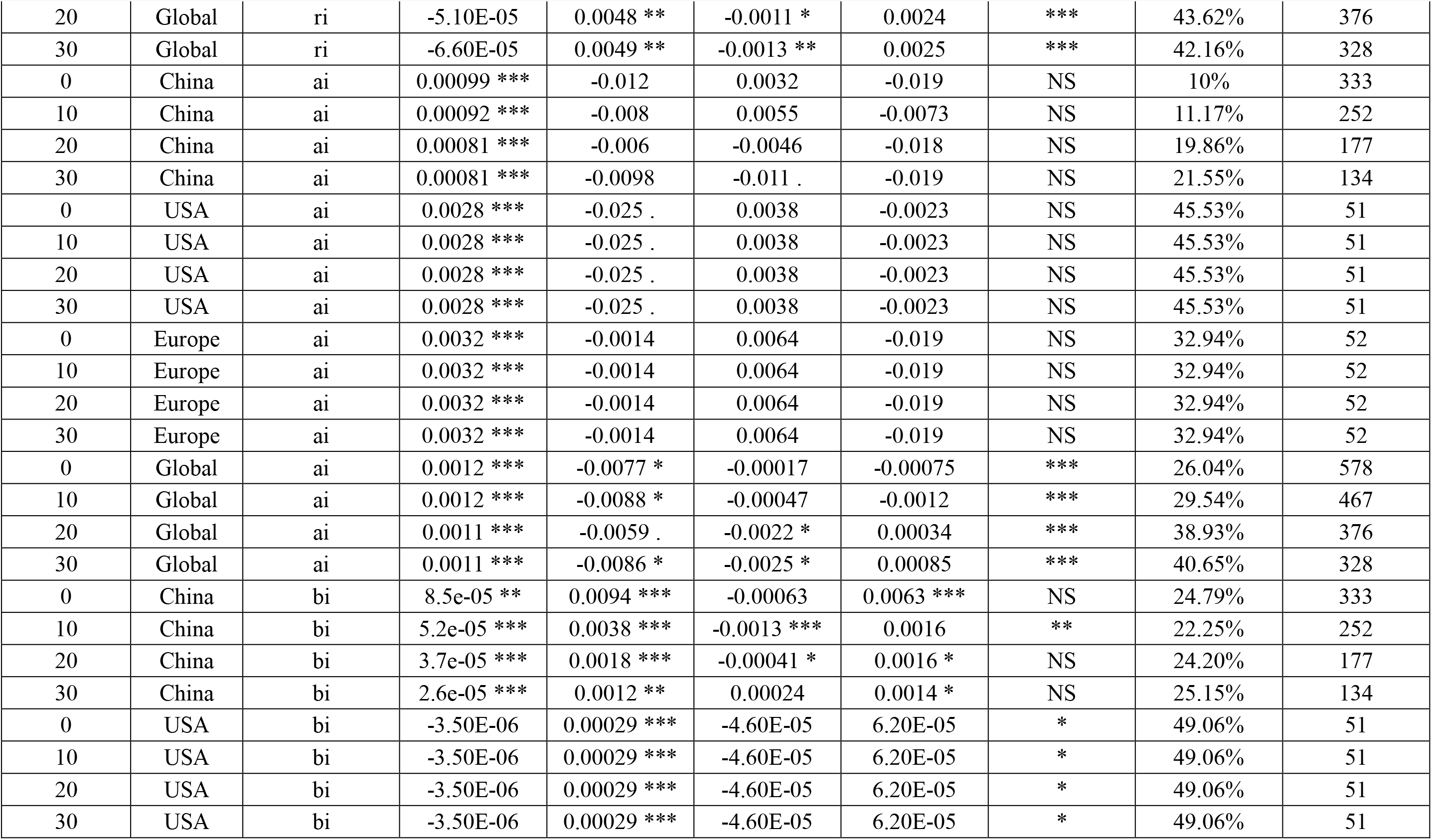
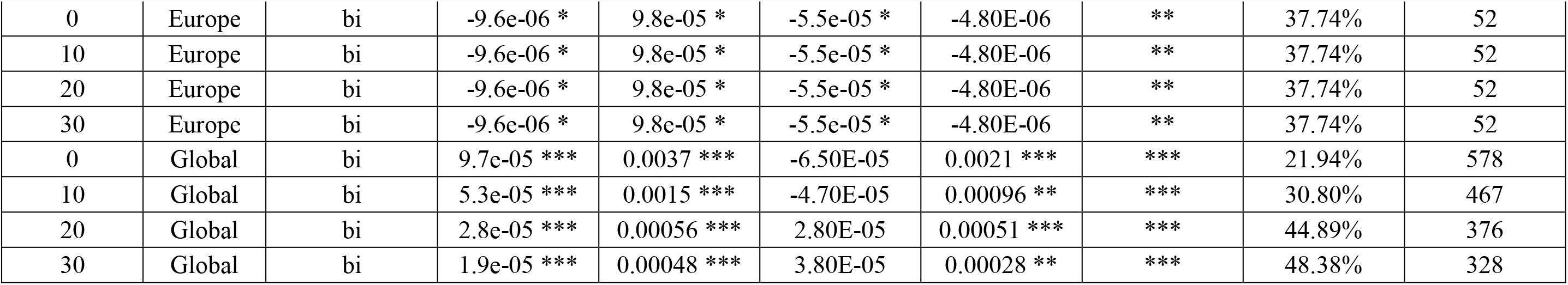
Associations of average daily increase rate (*r*_i_) of cumulative cases, maximum daily increase rate (*a*_i_), control efficiency (*b*_i_), of COVID-19 with founding population size during the initial 7 days after the first patient (*F*_i_), humanpopulation, climate factors (temperature and precipitation), and spatial autocorrelation based on analyses using Equation 2 (linear model) for locations with more than 0/10/20/30 cases. Bold values indicated the coefficients represent the significant effects (*p* < 0.05). + denotes the significant effects of spatial autocorrelation or temperature (*p* < 0.05), ns denotes non-significant effects. NA denotes not available (* *p* < 0.05, ** *p* < 0.01, *** P <0.001).

**Table S3:**
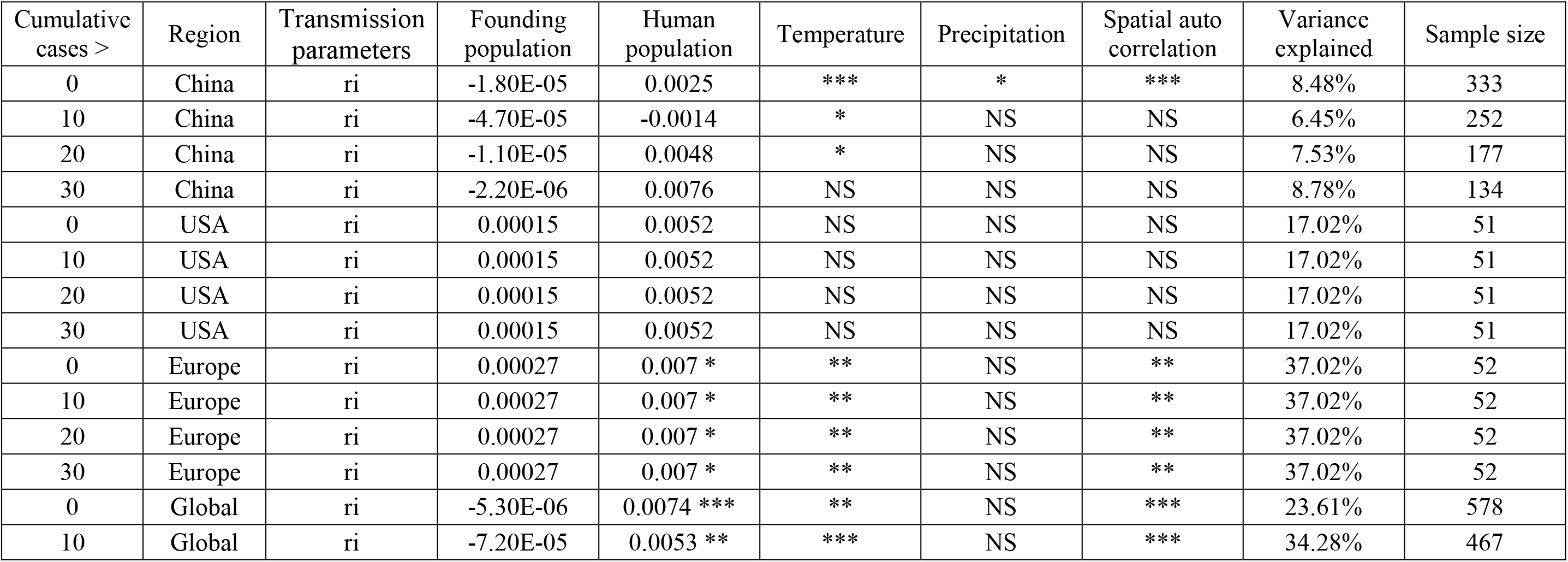
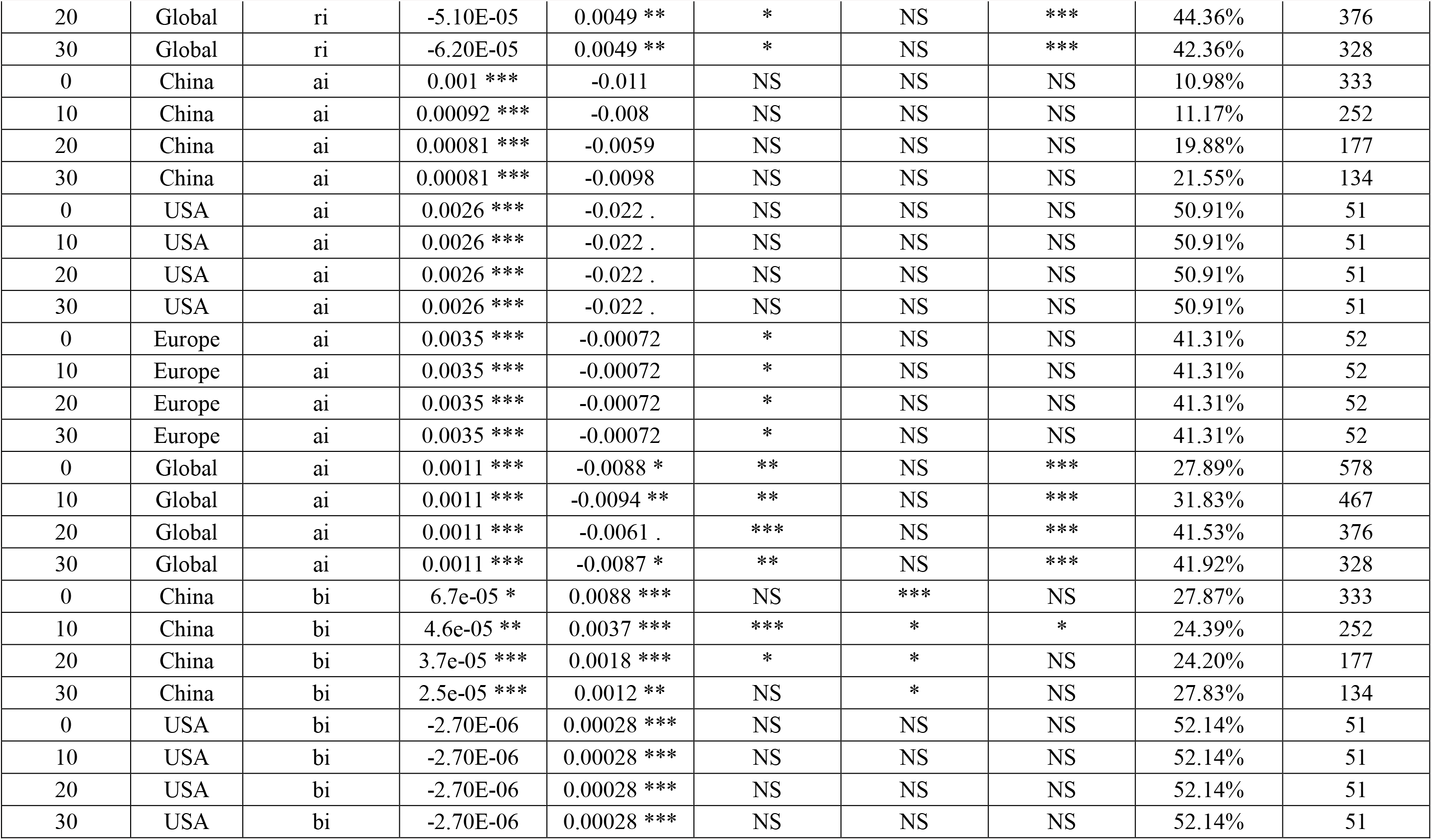
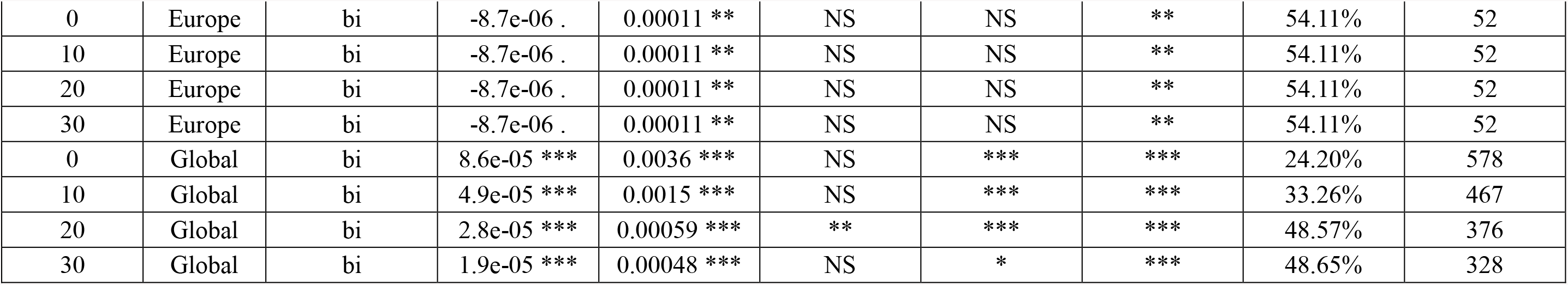
Associations of average daily increase rate (*r*_i_) of cumulative cases, maximum daily increase rate (*a*_i_), control efficiency (*b*_i_) of COVID-19 with founding population size during the initial 7 days after the first patient (*F*_i_), human population, climate factors(temperature and precipitation), and spatial autocorrelation based on analyses using Equation 3 (non-linear model) for locations with more than 0/10/20/30 cases. Bold values indicated the coefficients represent the significant effects (*p* < 0.05). + denotes the significant effects of spatial autocorrelation or temperature (*p* < 0.05), ns denotes non-significant effects. NA denotes not available (* *p* < 0.05, ** *p* < 0.01, *** *p*
0.001).

**Table S4:**
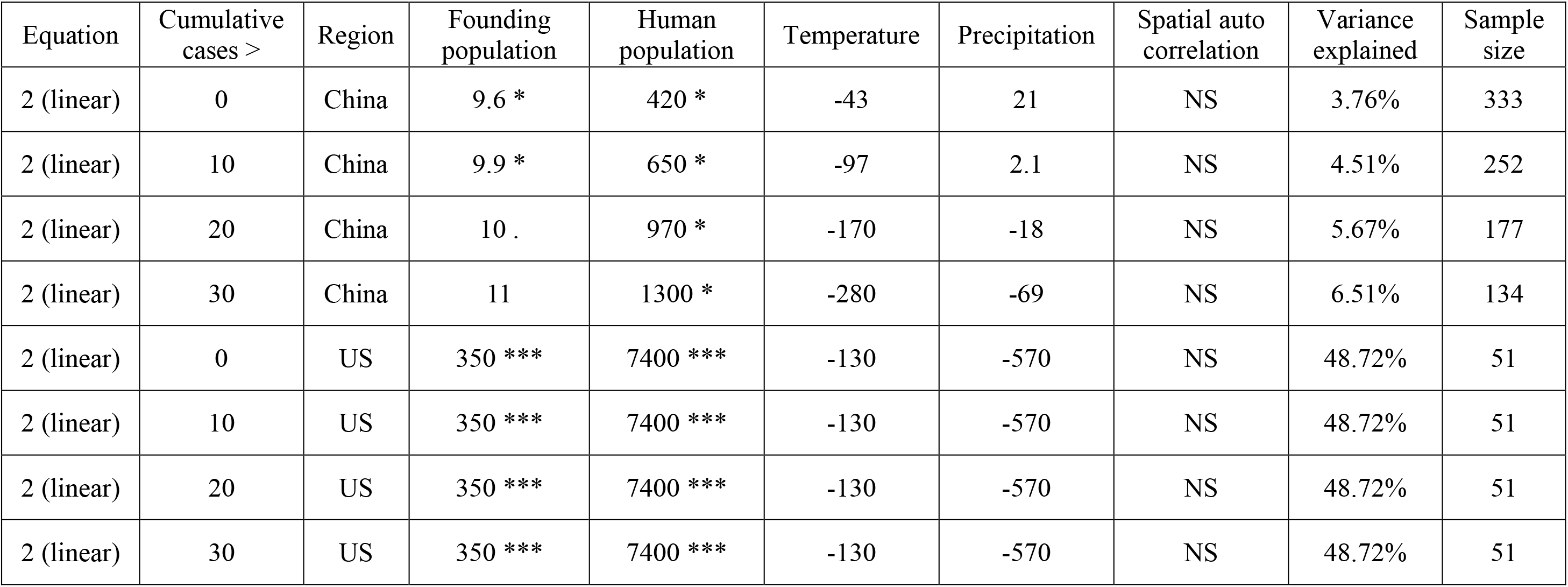
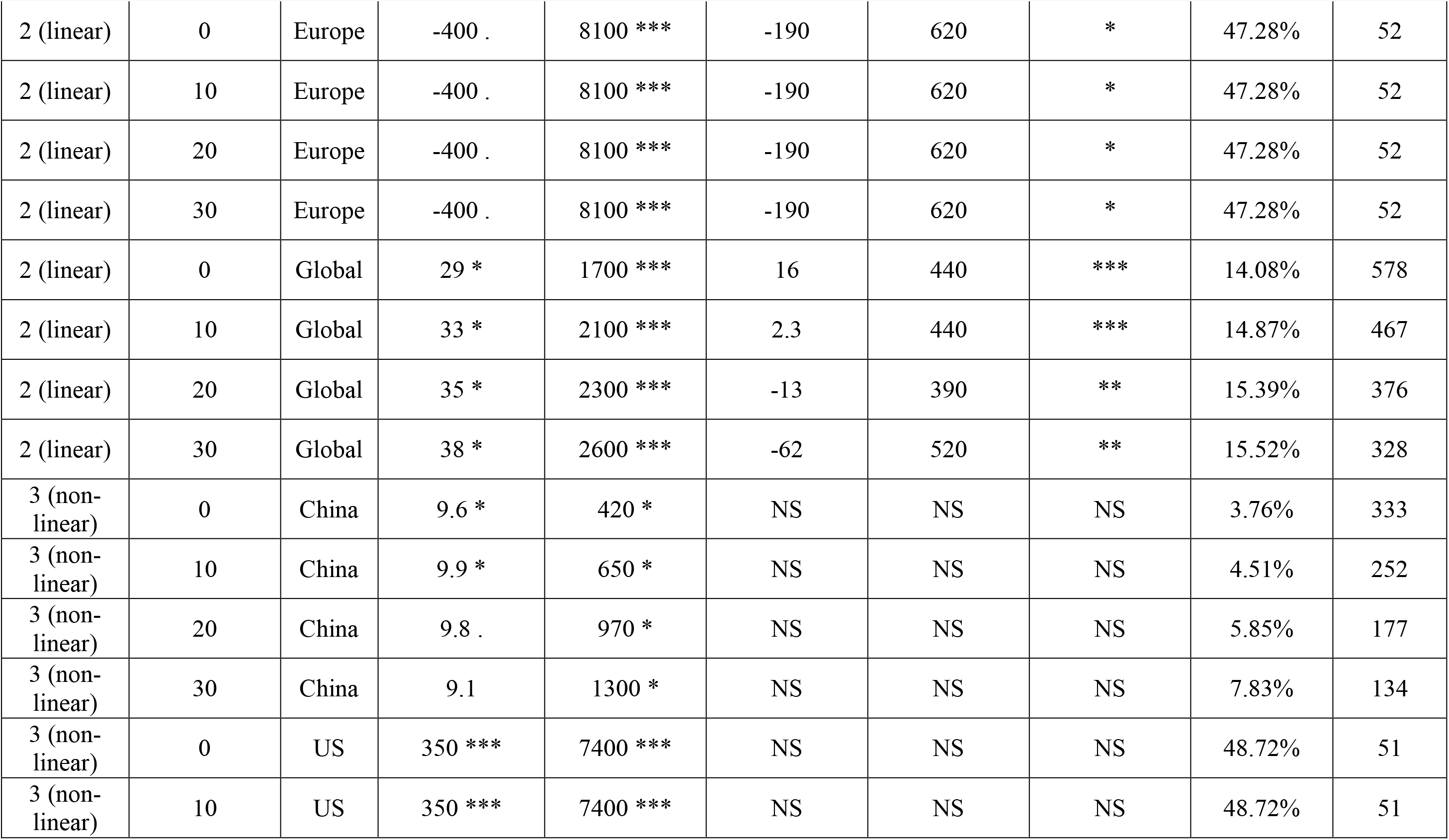
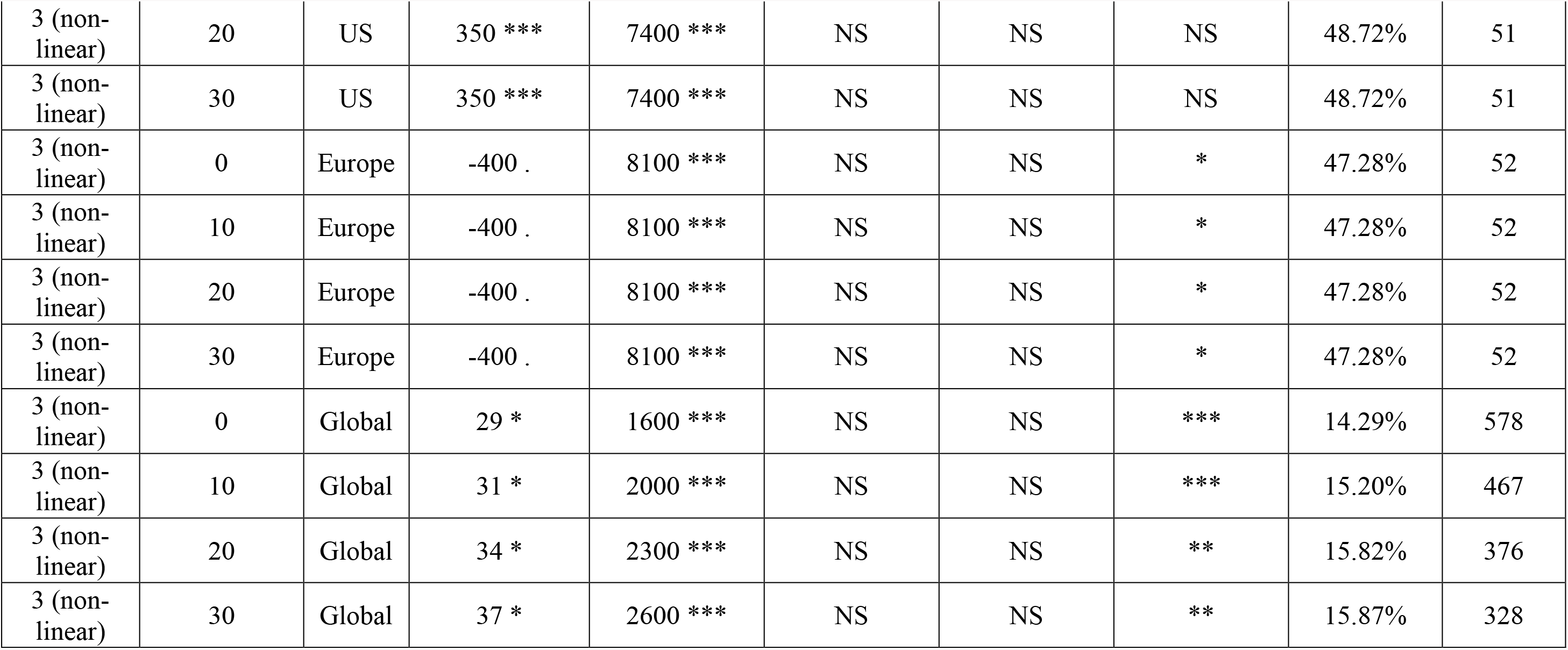
Associations of the number of cumulative cases (N) of COVID-19 with founding population size during the initial 7 days after the first patient (*F*_i_), human population, climate factors (temperature and precipitation), and spatial autocorrelation based on analyses using Equation 2 (linear model) and 3 (non-linear model) for locations with more than 0/10/20/30 cases. Bold values indicated the coefficients represent the significant effects (*p* < 0.05). + denotes the significant effects of spatial autocorrelation or temperature (*p* < 0.05), ns denotes non-significant effects. NA denotes not available (* *p* < 0.05, ** *p* < 0.01, *** *p* < 0.001).

